# Visualizing Geospatial and Temporal Phenotype Prevalences

**DOI:** 10.1101/2024.11.01.24316603

**Authors:** Michael Murray, Yuk-Lam Ho, Vidul A. Panickan, David Heise, Keith Connatser, Sumitra Muralidhar, Jacqueline Honerlaw, Kelly Cho

**Affiliations:** VA Boston Healthcare System, Boston, MA; Oak Ridge National Laboratory, Oak Ridge, TN; Office of Research and Development, VHA, Washington, D.C.; Harvard Medical School, Boston, MA

## Abstract

High-throughput phenotyping strategies are capable of classifying large volumes of patients. However, translating this data to real world applications is challenging. We have developed GeoPheno, a tool which displays the geospatial prevalences of EHR-based phenotypes in the Veteran population over time. Our flexible tool can display data from a wide array of phenotypes and is integrated with the CIPHER phenotype library, allowing users to view the definitions of the conditions being visualized.

## Introduction

Researchers utilize visualization tools to understand and communicate large scale data. Spatial maps are commonly used for depicting geospatial variations of disease burden and trends over time. While many mapping applications exist, a majority of these tools focus on a specific disease domain or are only used for single data source. This trend can be observed in the Centers for Disease Control’s (CDC) focused visualization tools and the wide array of COVID-19 tracking maps^1,2^.

However, viewing geospatial and temporal prevalences for multiple disease domains provides additional benefits to health data users. Public health officials are able to view and compare the burden of disease for a wide spectrum of conditions and researchers can gain a deeper understanding of cohort characteristics and use prevalences for hypothesis generation. GeoPheno is a new, interactive geospatial prevalence tool hosted by the Centralized Interactive Phenomics Resource (CIPHER), which is able to display prevalence data from conditions in any domain.

This brief report describes the GeoPheno tool design and applications for health research.

## Design

CIPHER is a publicly accessible platform developed by the US Department of Veterans Affairs (VA) and Oak Ridge National Laboratory (ORNL) to facilitate the development and reuse of computable phenotype definitions using health data (Figure 1).^3^ The platform contains a knowledgebase of phenotype definition metadata and phenotyping resources. CIPHER works with collaborators to create tools related to phenotype development and knowledge extraction, integrate tools into the platform, and link them to related definitions in the phenotype library.

**Figure 1.**
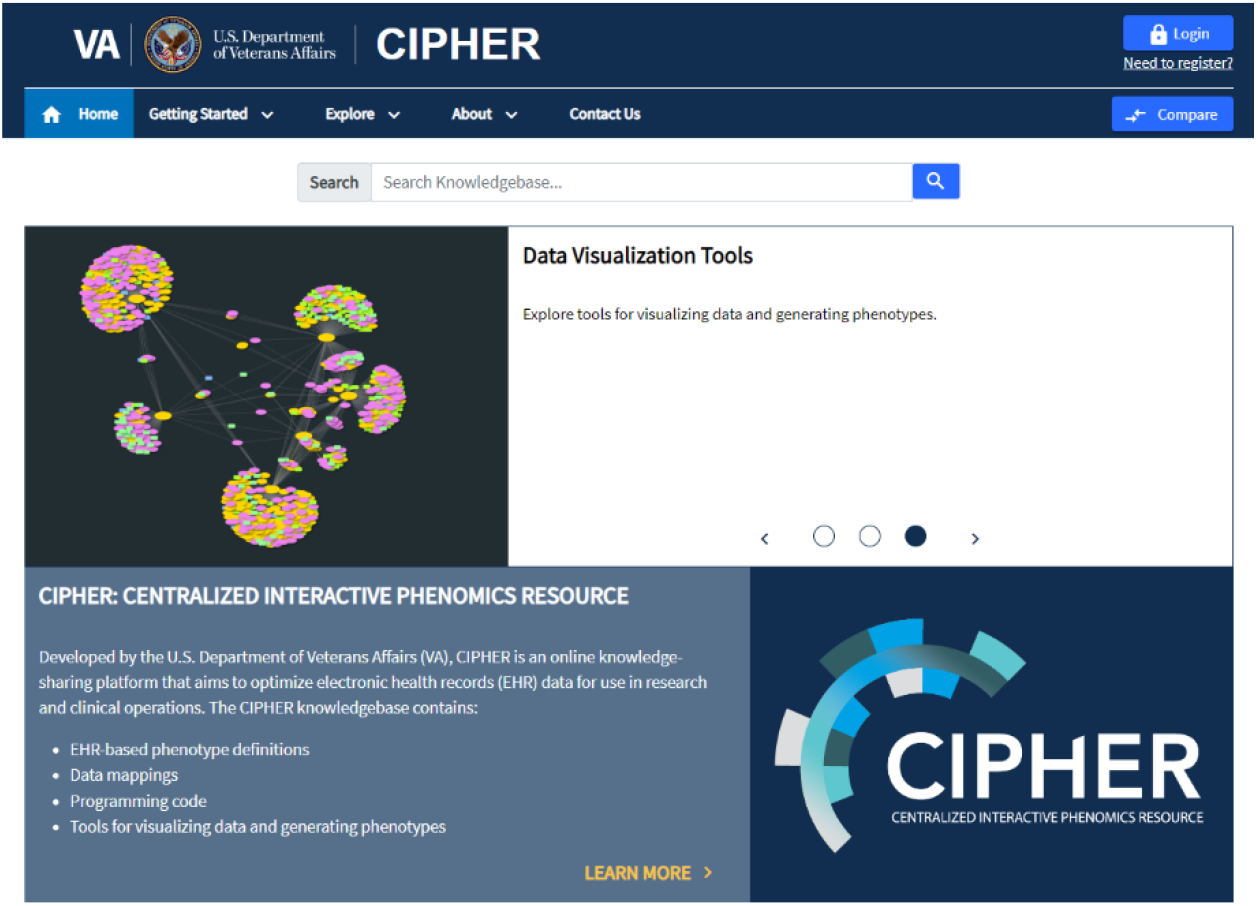
CIPHER’s home page

The GeoPheno tool was developed for CIPHER with support from the Center for Learning Health System (CELEHS) at Harvard Medical School and the Predictive Analytics Research Solution and Execution (PARSE). The tool is built on RShiny, a framework for deploying visualizations of disease prevalence data over United States in R.

The tool offers a wide array of features to help users better understand conditions of interest. Researchers can utilize the collapsible explorer panel on the left side of the tool to select a phenotype of interest and create figures using available options. First, a dropdown allows the selection of source of data, followed by selecting an available phenotype within that source. Doing so populates a national map, seen on the top of the screen, with average prevalences in percentages based on available data for the selected phenotype (Figure 2). The color gradient displayed on the map is determined by quintiles of the overall percentages. A user may hover over states to view the total number of cases and average prevalence for each state. Users can also move a slider to define year range of interest. The map display will automatically adjust to the average prevalence of the selected years. Currently, U.S. states are the only geographic option, but future versions may expand to include additional geographic boundaries, such as including VA Veterans Integrated Services Networks (VISNs).

**Figure 2.**
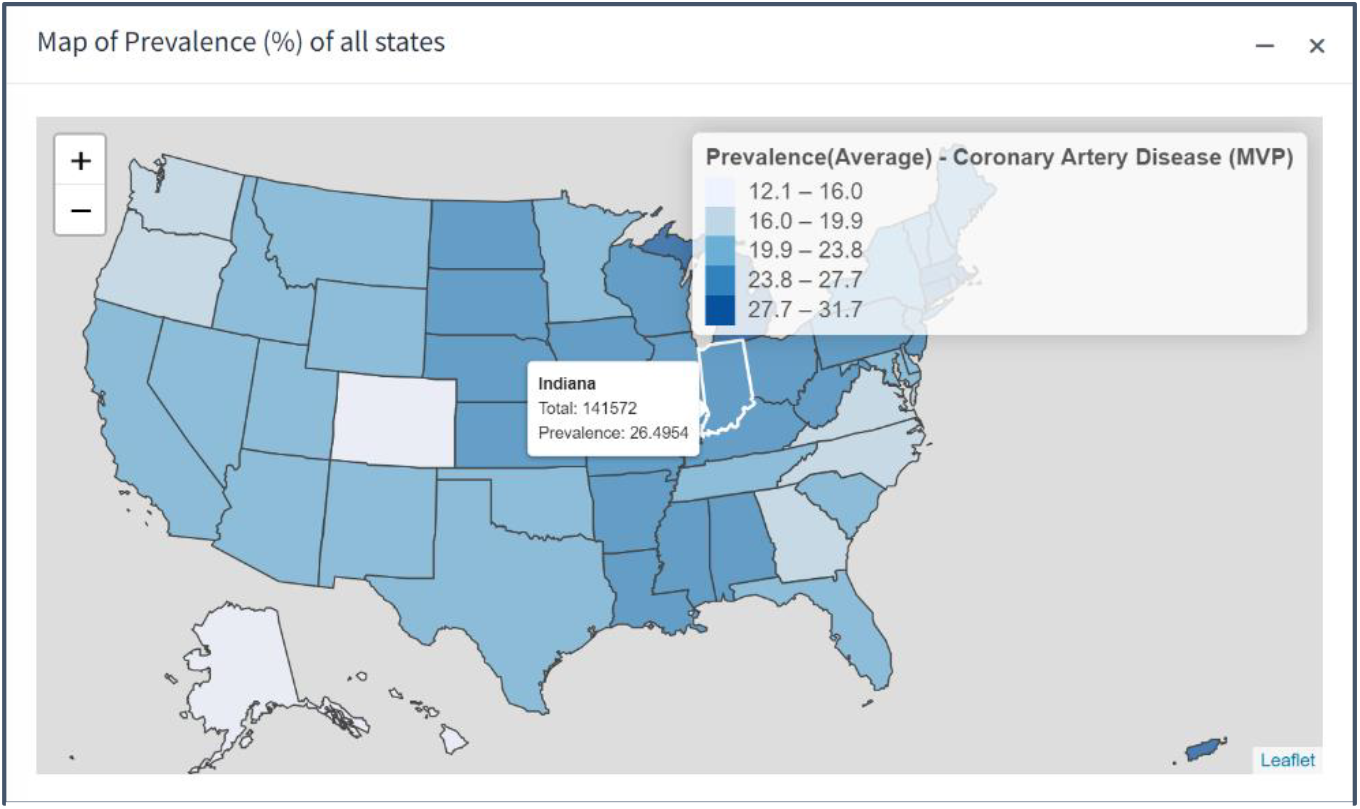
GeoPheno displaying a prevalence map for Coronary Artery Disease.

Below the map, there are several plot options to allow users to explore time trends and summary statistics by selecting state options (Figure 3). User can use dropdown menu to select a state or states of interest. Upon selection, GeoPheno will populate a line plot showing prevalence trends over time by year, and bar plots breaken down by age, race, and sex. These categories are not comprehensive. The groupings for race include White, Black, Asian, American Indian, Native Hawaiian or Pacific Islander, and other; options for sex are male and female; patient age is broken down by ten-year windows, with age groups below 40 and above 90 collapsed. These plots can be downloaded as image files for further use. At the bottom, a table view is available for users to see prevalences for all states as well as a summary statistics breakdown by demographics for the selected phenotype. All cells with counts below 11 in the table are replaced with – 999 to reduce risk of patient re-identification.

**Figure 3.**
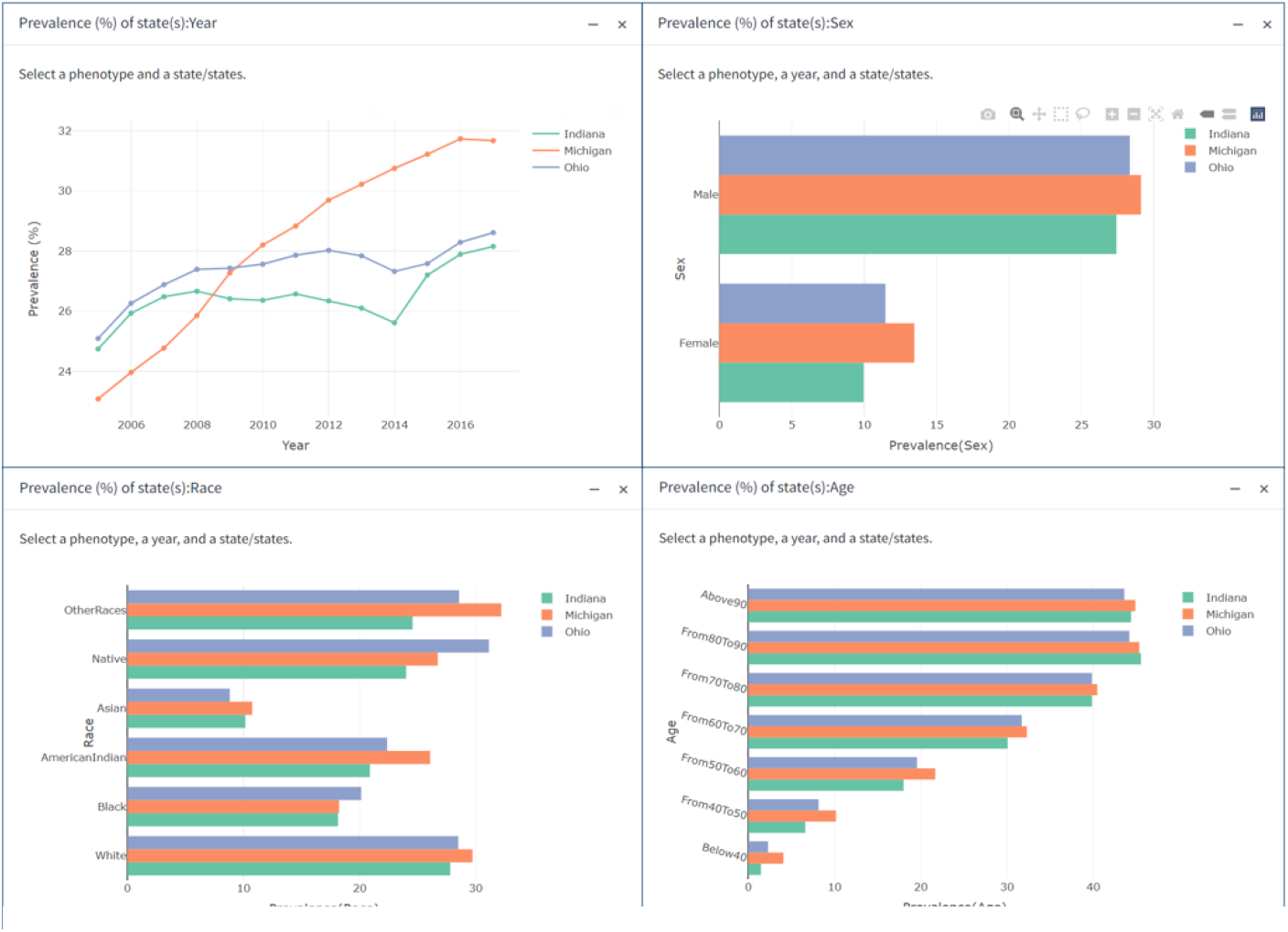
Charts showing prevalence in three states over time, as well as breakdowns by age, sex, and race. Users can download images of these charts.

One of GeoPheno’s most significant features is the bidirectional connection between applied prevalence data in the tool and the phenotype definition in CIPHER. A user may click a button below the phenotype selected to be brought directly to the definition in the phenotype library. This definition contains all of the metadata required to replicate the phenotype in a different population using the CIPHER standard, as well as references and citations.^4^ A link to GeoPheno will appear on phenotype pages in CIPHER that have prevalence data in the tool. The linkage is accomplished by coordinating unique identifiers between both sets of data. When a link between the two is clicked, the receiving application gets the ID of the phenotype to open as a query string parameter.

## Discussion

The GeoPheno tool enables display of prevalences cataloged within the CIPHER phenotype library and allows comparison of multiple phenotypes across institutions. Initial set of phenotypes shown in the tool include a few selected phenotypes catalogued within CIPHER and applied to the VA population to generate this summary level data. Additionally, an equivalent set of conditions collected by US Behavioral Risk Factor Surveillance System (BRFSS) survey have been included in the first release of the tool. A major set of phenotypes collected by CIPHER, including phenotypes developed using high-throughput multimodal automated phenotyping (MAP) unsupervised classification algorithm applied to all VHA patients and phenotypes published in the Million Veteran Program (MVP) genome-wide phenome-wide association study (gwPheWAS), are being finalized and reviewed for addition to GeoPheno in upcoming releases.^5,6^ While the initial release focus on the VHA population, we plan to incorporate phenotypes from many other data sources and CIPHER partners in future releases.

Additionally, we hope to include user submitted phenotypes. Researchers who have submitted phenotypes to CIPHER can choose to contact us to submit prevalence data for inclusion in GeoPheno. By incorporating data from a wider range of populations and linking these prevalences to their definitions in CIPHER, we will continue to expand the utility of this tool for phenotype comparison and selection, as well as hypothesis generation. GeoPheno can be found at https://phenomics.va.ornl.gov/geopheno. CIPHER can be found at https://phenomics.va.ornl.gov/.

## Data Availability

The phenotype definition metadata described in this article will be made available on
https://phenomics.va.ornl.gov .

## References

1. Center for Disease Control. Interactive Web Applications & Data [Internet]. 2020 [Cited 2024 March]. Available from: https://www.cdc.gov/gis/interactive-applications.htm

2. Saran, S., Singh, P., Kumar, V. et al. Review of geospatial technology for infectious disease surveillance: use case on COVID-19. J Indian Soc Remote Sens 48, 1121–1138 (2020). doi:10.1007/s12524-020-01140-5

3. Honerlaw J, Ho YL, Fontin F, et al. Centralized Interactive Phenomics Resource: an integrated online phenomics knowledgebase for health data users. J Am Med Inform Assoc. Published online March 13, 2024. doi:10.1093/jamia/ocae042

4. Honerlaw J, Ho YL, Fontin F, et al. Framework of the Centralized Interactive Phenomics Resource (CIPHER) standard for electronic health data-based phenomics knowledgebase. J Am Med Inform Assoc. 2023;30(5):958–964. doi:10.1093/jamia/ocad030

5. Liao KP, Sun J, Cai TA, et al. High-throughput multimodal automated phenotyping (MAP) with application to PheWAS. J Am Med Inform Assoc. 2019;26(11):1255–1262. doi:10.1093/jamia/ocz066

6. Verma A, Huffman JE, Rodriguez A, et al. Diversity and scale: Genetic architecture of 2068 traits in the VA Million Veteran Program. Science. 2024;385(6706):eadj1182. doi:10.1126/science.adj1182

